# Cross-reactive Bundibugyo antibody responses after licensed Ebola vaccines

**DOI:** 10.64898/2026.05.27.26354223

**Authors:** Edouard Lhomme, Aurélie Wiedemann, Ahidjo Ayouba, Safaa Ben-Farhat, Guillaume Thaurignac, Céline Roy, Abdoul Habib Beavogui, Seydou Doumbia, Mark Kieh, Bailah Leigh, Samba Sow, Stephen A Migueles, Deborah Watson-Jones, Yazdan Yazdanpanah, Rodolphe Thiébaut, Martine Peeters, Laura Richert, Yves Levy, PREVAC study Team

## Abstract

**Background:** The ongoing Bundibugyo virus disease (BDBV) outbreak in Central Africa highlights the absence of approved vaccines specifically targeting BDBV. Whether licensed Zaire ebolavirus (EBOV) vaccines induce cross-reactive immunity against BDBV remains largely unknown.

**Methods:** We performed an immunogenicity analysis using serum samples from participants enrolled in the PREVAC randomized clinical trial evaluating licensed Ebola vaccine strategies in West Africa. Samples collected at day 28 (D28) and month 3 (M3) following vaccination with rVSVΔG-ZEBOV-GP or Ad26.ZEBOV/MVA-BN-Filo were assessed using a multiplex Luminex assay against glycoproteins from multiple filoviruses, including EBOV Kikwit, EBOV Mayinga, BDBV, Sudan virus, Reston virus, and Marburg virus.

**Results:** A total of 179 samples were analysed. Detectable cross-reactive antibody responses against BDBV were observed across vaccine groups, timepoints, and age categories. However, BDBV responses remained substantially lower than homologous EBOV responses. In rVSV recipients, median BDBV responses (net MFI) reached 282 (IQR 164–644) at D28 compared with 1788 (832–3311) against the homologous Kikwit antigen. Similar patterns were observed following rVSV booster vaccination and Ad26.ZEBOV/MVA-BN-Filo vaccination. The heterologous Ad26/MVA regimen demonstrated increasing BDBV responses between D28 and M3.

**Conclusions:** Licensed EBOV vaccines induced detectable but quantitatively reduced cross-reactive antibody responses against BDBV. Although no direct assessment of vaccine efficacy against BDBV disease was possible, these findings support the plausibility of partial heterologous immunity following EBOV vaccination. In the absence of approved BDBV-specific vaccines, these data support the urgent evaluation of currently available Ebola vaccines during BDBV outbreaks and reinforce the importance of developing broadly protective pan-filovirus vaccines.

## Introduction

The ongoing outbreak of Ebola virus disease caused by Bundibugyo virus (BDBV) in the Democratic Republic of the Congo (DRC) and Uganda has rapidly escalated as a major global health concern, with 861 reported cases and an estimated case fatality rate of approximately 21% as of May 25, 2026.^1^ On May 16, 2026, the World Health Organization declared the epidemic a Public Health Emergency of International Concern following evidence of sustained transmission across multiple health zones, cross-border spread, and increasing suspected BVBD-associated mortality.^2^

BDBV is a relatively understudied ebolavirus species that was responsible for only two previous documented outbreaks since its identification in 2007. In contrast to Zaire ebolavirus (EBOV), no vaccine or specific therapeutics are currently available for the prevention or treatment of BDBV infection, leaving populations at risk without proven medicinal countermeasures in the context of ongoing transmission. Moreover, current diagnostics that can be used in decentralized settings do not detect BDBV strains, thus delaying thus significantly rapid implementation of countermeasures to interrupt transmission chains. Genomic analyses from the current outbreak suggest a zoonotic spillover event and confirm genetic divergence from EBOV strains targeted by licensed vaccines.^3^ Therefore, whether currently licensed EBOV vaccines may induce cross reactive immunity against BDBV remains debatable. Preclinical data have suggested partial and inconsistent heterologous protection with the rVSV-ZEBOV platform,^4–6^ whereas evidence is scarce for the Ad26.ZEBOV/MVA-BN-Filo regimen.^7^ Finally, immunogenicity data against BDBV in humans are extremely limited.

In this context, we assessed cross-reactive antibody responses against multiple ebolavirus species, including BDBV, using serum samples from participants enrolled in the PREVAC randomised clinical trial that evaluated the rVSV-ZEBOV (Ervebo®, MSD), and Ad26.ZEBOV/MVA-BN-Filo (Zabdeno®, Mvabea®, Janssen), vaccine strategies in adults and in children in West Africa,^8^ to provide urgently needed insights into potential cross-protective immunity.

## Methods

PREVAC (Partnership for Research on Ebola Vaccinations) was a multicentre, randomised, placebo-controlled phase 2 trial conducted in Guinea, Liberia, Mali, and Sierra Leone to evaluate the safety and immunogenicity of three Ebola vaccine strategies in healthy adults and children (NCT02876328). Participants were randomly assigned to receive either Ad26.ZEBOV (0.5□ml; 5□×□10^10^ viral particles) followed by MVA-BN-Filo (0.5□ml; 1□×□10^8^ infectious units) 56 days later (the Ad26–MVA group), rVSVΔG-ZEBOV-GP (1.0□ml; 9.4□×□10^7^ plaque-forming units) followed by placebo 56 days later (the rVSV group), and rVSVΔG-ZEBOV-GP followed by rVSVΔG-ZEBOV-GP 56 days later (the rVSV–booster group), or placebo. Detailed trial methods and primary results have been previously reported.^8,9^

For this analysis of cross-reactive antibody responses against ebolavirus species, we analysed a subset of available serum samples collected from vaccinated participants at day 28 (D28) and month 3 (M3) post-vaccination corresponding to expected peak time points of vaccine immune responses. Samples were selected on the basis of availability at the biobank from the Institut de Recherche pour le Développement (IRD, Montpellier, France) laboratory and included both adults and children across vaccine groups (Table 1).

**Table 1.** Characteristics of the study population included in the study from the PREVAC trial.

| Characteristics | rVSV Group |  | rVSV Booster group |  | Ad26 / MVA Group |  |
| --- | --- | --- | --- | --- | --- | --- |
|  | D28<br>(n = 28) | M3<br>(n = 44) | D28<br>(n = 10) | M3<br>(n = 25) | D28<br>(n = 17) | M3<br>(n = 55) |
| **Age** |  |  |  |  |  |  |
| Adult | 12 (42.9%) | 31 (70.5%) | 6 (60.0%) | 16 (64.0%) | 7 (41.2%) | 36 (65.5%) |
| Children | 16 (57.1%) | 13 (29.5%) | 4 (40.0%) | 9 (36.0%) | 10 (58.8%) | 19 (34.5%) |
| 1-4 yr | 4 (9.1%) | 1 (10.0%) | 3 (12.0%) | 4 (23.5%) | 7 (12.7%) | 3 (12.0%) |
| 5-11 yr | 6 (13.6%) | 1 (10.0%) | 3 (12.0%) | 3 (17.6%) | 6 (10.9%) | 3 (12.0%) |
| 12-17 yr | 3 (6.8%) | 2 (20.0%) | 3 (12.0%) | 3 (17.6%) | 6 (10.9%) | 3 (12.0%) |
| **Study site (Country)** |  |  |  |  |  |  |
| CVD (Mali) | 5 (17.9%) | 2 (4.5%) | 3 (30.0%) | 3 (12.0%) | 8 (47.1%) | 3 (5.5%) |
| Landreah (Guinea) | 2 (7.1%) | 28 (63.6%) | 1 (10.0%) | 12 (48.0%) | 1 (5.9%) | 28 (50.9%) |
| Maferinyah (Guinea) | 0 (0%) | 0 (0%) | 2 (20.0%) | 1 (4.0%) | 0 (0%) | 5 (9.1%) |
| Mambolo (Sierra Leone) | 5 (17.9%) | 1 (2.3%) | 1 (10.0%) | 4 (16.0%) | 0 (0%) | 8 (14.5%) |
| Redemption (Liberia) | 9 (32.1%) | 7 (15.9%) | 2 (20.0%) | 4 (16.0%) | 6 (35.3%) | 8 (14.5%) |
| UCRC (Mali) | 7 (25.0%) | 6 (13.6%) | 1 (10.0%) | 1 (4.0%) | 2 (11.8%) | 3 (5.5%) |

CVD: Center for Vaccine Development (Mali) ; UCRC: University Clinical Research Center (Mali)

Cross-reactive antibody responses were assessed using a multiplex bead-based Luminex assay. Recombinant glycoprotein antigens representing six filoviruses were evaluated, including EBOV Mayinga (MAY), EBOV Kikwit (KIK), BDBV virus (Uganda 2007), Sudan virus (Gulu), Reston virus, and Marburg virus (Musoke 1980). Plasma samples were tested at a working dilution of 1:200 in duplicate. Samples were incubated with antigen-coated beads for 16 hours at 4°C under agitation and protected from light, followed by sequential incubation with biotinylated mouse anti-human antibodies and streptavidin-phycoerythrin. Median fluorescence intensity (MFI) was acquired on a Luminex INTELLIFLEX platform. Duplicate measurements with coefficients of variation greater than 20% were repeated (*Ayouba JCM 2016, Berry BioRxiv 2026*).^10,11^ The median of the duplicates per sample was used in the statistical analysis. Antibody responses were analysed descriptively across vaccine groups, age strata and timepoints.

## Results

A total of 179 serum samples from vaccinated participants enrolled in the PREVAC trial were included. Samples were collected at D28 (n=55, 30.7%) and M3 (n=124, 69.3%) following vaccination. Participants received the heterologous Ad26/MVA regimen (72 samples, 40.2%), a single-dose rVSV vaccine (72 samples, 40.2%), or a rVSV booster strategy (35 samples, 19.6%). Adults (n=108, 60.3%) and children (n=71, 39.7%) samples from multiple PREVAC study sites in West Africa (Table 1) were included. Overall, participant samples reflect the multicentre and age-diverse PREVAC trial population^8^ enabling exploratory comparisons across vaccine platforms, age strata, and timepoints.

Cross-reactive antibody responses against EBOV (KIK and MAY strains) and BDBV were detected across all vaccine strategies and timepoints at both D28 and M3 (Figure 1, Figure S1) while responses against Marburg were below the limit of detection. This pattern was consistent across adults and children and showed heterologous humoral immunity against BDBV following EBOV vaccination with either the rVSV vaccine or the Ad26.ZEBOV/MVA-BN-Filo vaccine (Table 2, Figure 1). BDBV-specific responses were consistently lower than homologous EBOV responses across all vaccine regimens and timepoints. In rVSV recipients, median responses against BDBV were 282 (IQR 164–644) at D28 and 130 (106–436) at M3, compared with 1788 (832–3311) and 1069 (747–1876) against EBOV Kikwit, respectively. Similar findings were observed in the rVSV booster group (BDBV: 236 [114–865] at D28 and 246 [137–553] at M3), remaining substantially lower than homologous Kikwit responses.

**Table 2.** Description of the antibody response at Day 28 and Month 3 post-vaccination by rVSV, rVSV-rVSV and Ad26/MVA respectively.

| Vaccine arm | Filovirus species and variants * |  |  |  |  |  |
| --- | --- | --- | --- | --- | --- | --- |
|  | EBOV (Mayinga) | EBOV (Kikwit) | SUDV | BDBV | RESTV | MARV |
| **rVSV Group** |  |  |  |  |  |  |
| D28 (*n* = 28) | 4289 (2693, 7567) | 1788 (832, 3311) | 1512 (654, 2344) | 282 (164, 644) | 42 (27, 68) | 12 (6, 31) |
| M3 (*n* = 44) | 2086 (1245.25, 2914) | 1069 (747, 1876) | 1029 (302, 2035) | 131 (106, 436) | 34 (24, 50) | 10 (6, 24) |
| **rVSV Booster Group** |  |  |  |  |  |  |
| D28 (*n* = 10) | 3503 (2442, 4365) | 819 (336, 1454) | 1237 (363, 2256) | 236 (114, 865) | 27 (16, 41) | 17 (8, 20) |
| M3 (*n* = 25) | 5274 (4174, 7841) | 2648 (1750, 3653) | 875 (276, 1662) | 246 (137, 553) | 80 (58, 107) | 14 (5, 32) |
| **Ad26 / MVA Group** |  |  |  |  |  |  |
| D28 (*n* = 17) | 924 (522, 2404) | 184 (118, 601) | 518 (231, 1351) | 83 (44, 131) | 17 (11, 24) | 8 (5, 14) |
| M3 (*n* = 55) | 6311 (4309, 8775) | 2602 (1472, 3782) | 1991 (968, 3522) | 433 (236, 994) | 136 (75, 354) | 49 (17, 101) |

\* Median (Q1, Q3)
EBOV, Ebolavirus; SUDV, Sudanvirus; BDBV, Bundibunyovirus; RESTV, Restonvirus; MARV, Marburgvirus

**Figure 1.**
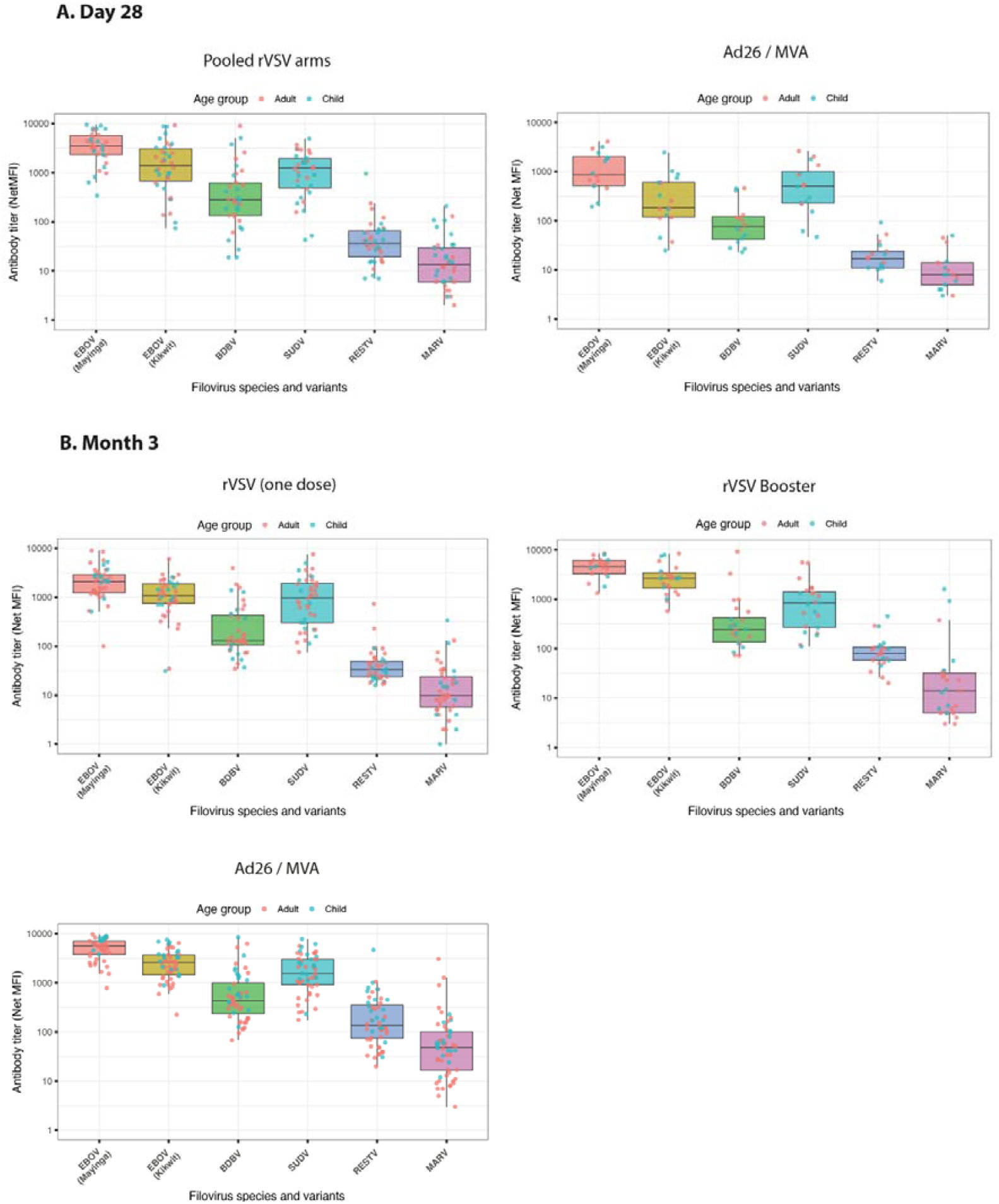
Description of the binding antibody response measured by multiplex Luminex (net MFI) at day 28 and Month 3 after rVSV, rVSV booster and Ad26 / MVA vaccination, respectively.

Following the heterologous Ad26.ZEBOV/MVA-BN-Filo regimen, BDBV responses were higher at M3 (median MFI 433 (236–994)) than at D28 (median MFI 83 (44–131)), but remained markedly lower than corresponding EBOV Kikwit responses. These findings show that current licensed EBOV vaccines induced limited but measurable heterologous humoral responses against BDBV.

## Discussion

The ongoing BDBV outbreak in the DRC and Uganda is occurring in a context of extreme vulnerability, with no licensed vaccines or therapeutics specifically available for this ebolavirus species. Therefore, evidence supporting the ability of existing vaccines developed against Zaire EBOV to induce a humoral response and a degree of cross-protective immunity for BDBV is of immediate public health relevance.

In this analysis, we showed that both licensed EBOV vaccines induced detectable cross-reactive binding antibody responses against BDBV glycoprotein. These responses were observed across vaccine platforms, age groups, and timepoints, indicating heterologous humoral recognition following EBOV-based vaccination. Responses to BDBV were consistently and substantially lower than those against homologous EBOV glycoproteins (Kikwit and Mayinga), indicating limited quantitative cross-reactivity, but were markedly higher than responses against Marburg virus GP. We did not find a meaningful increase in BDBV responses following the homologous rVSV boost consistent with prior findings for homologous EBOV responses in PREVAC trial.^8^ These observations align with non-human primate data showing partial and incomplete protection against BDBV following rVSV-based vaccination.^5^ By contrast, the heterologous Ad26.ZEBOV/MVA-BN-Filo regimen showed a progressive increase in BDBV-specific antibodies between day 28 and month 3, suggesting delayed maturation of cross-reactive humoral responses.

Of note, the absence of a validated immunological correlate of protection or a minimal threshold of protection for anti-EBOV GP IgG antibodies may limit the interpretation of these results. However, EBOV GP-binding antibodies have been widely used as immunological readouts in vaccine studies and these measures have previously shown associations with vaccine-induced protection in EBOV studies^12–14^ supporting their use as biological relevant measures in the present study. Moreover, the Luminex platform used here has previously demonstrated concordance with the reference FANG ELISA assay.^11^

Several limitations warrant consideration. This exploratory analysis was performed on a subset of available PREVAC samples rather than on the full trial population. Only binding antibody responses were assessed, without measurement of neutralisation or cellular immunity. Finally, antigenic differences between the 2007 Uganda BDBV variant used in this study and viruses circulating in the current outbreak may limit direct extrapolation. However, this limitation could be applied also when licensed vaccines (including the older EBOV variants Mayinga and Kikwit) are deployed during recent EBOV outbreaks.

Although no direct assessment of vaccine efficacy against BDBV infection or disease can be made, in the context of an unfolding outbreak and the absence of BDBV-specific medical countermeasures, these data provide biological support for the the plausibility of partial heterologous immunity against BDBV following EBOV vaccination.

In the absence of approved BDBV-specific vaccines or therapeutics, currently licensed EBOV vaccines remain among the only immediately deployable vaccine platforms available in affected regions. Importantly, the markedly reduced magnitude of BDBV responses related to homologous EBOV responses underscores the uncertainty regarding the level and durability of potential clinical protection. These findings highlight the urgent need for prospective clinical evaluation of currently available Ebola vaccines during BDBV outbreaks, including assessment of protection against infection and disease severity. More broadly, they reinforce the necessity of accelerating development of broadly protective or multivalent pan-filovirus vaccine strategies capable of spanning genetically diverse ebolavirus species.

### Notes

The members of the writing committee assume responsibility for the overall content and integrity of this article.

### Trial registration numbers

ClinicalTrials.gov number, NCT02876328 https://clinicaltrials.gov/study/NCT02876328

EudraCT number 2017-001798-18 https://www.clinicaltrialsregister.eu/ctr-search/trial/2017-001798-18/3rd

Pan African Clinical Trials Registry number, PACTR201712002760250.

### Open Science

PREVAC pseudonymized data are sensitive data collected through a clinical trial. Therefore, for participants data protection and privacy, legal and ethical reasons, the PREVAC pseudonymized data cannot be shared publicly.

The PREVAC metadata are available on the Recherche Data Gouv repository, DOI: 10.57745/PVXRMC (https://doi.org/10.57745/PVXRMC).

Authorization for the use of the PREVAC/PREVAC-UP pseudonymized clinical and immunological data must be granted by the sponsors and the concerned country PIs. The final decision to either authorize or not the use of the data and data transfer lies with the sponsors in order to ensure the data request is in accordance with legal and ethical regulations. If a data request is granted, a DTA (data transfer agreement) must be established.

For any additional question regarding access to the PREVAC/PREVAC UP pseudonymized data please contact.

## Data Availability

PREVAC pseudonymized data are sensitive data collected through a clinical trial. Therefore, for participants data protection and privacy, legal and ethical reasons, the PREVAC pseudonymized data cannot be shared publicly.
The PREVAC metadata are available on the Recherche Data Gouv repository, DOI: 10.57745/PVXRMC (https://doi.org/10.57745/PVXRMC).
Authorization for the use of the PREVAC/PREVAC-UP pseudonymized clinical and immunological data must be granted by the sponsors and the concerned country PIs. The final decision to either authorize or not the use of the data and data transfer lies with the sponsors in order to ensure the data request is in accordance with legal and ethical regulations. If a data request is granted, a DTA (data transfer agreement) must be established.
For any additional question regarding access to the PREVAC/PREVAC UP pseudonymized data please contact.

## Funding

This research was supported in part by the US National Institutes of Health (NIH), by the French Institut national de la santé et de la recherche médicale (Inserm) and by the London School of Hygiene and Tropical Medicine (LSHTM).

The clinical trial was conducted with the support of Janssen, Bavarian Nordic and Merck & Co., Inc., Rahway, NJ, USA who provided the vaccines according to the EBOVAC 1 grant agreement.

This project is part of the EDCTP2 programme supported by the European Union (grant number RIA2017S-2014 – PREVAC-UP).

This project has received funding from the Innovative Medicines Initiative 2 Joint Undertaking (IMI2JU) under grant agreement No 115854, EBOVAC1. This Joint Undertaking receives support from the European Union’s Horizon 2020 research and innovation program and EFPIA.

Funding provided in part by NCI contract HHSN261201500003I and 75N91019D00024 through the Frederick National Laboratory for Cancer Research. The content of this publication does not necessarily reflect the views or policies of the Department of Health and Human Services, nor does mention trade names, commercial products, or organizations imply endorsement by the U.S. Government.

The project has been funded by a dedicated Inserm allocation on behalf of the French Research Ministry.

The dissemination represents only the authors’ views and IMI2JU is not responsible for any use of the information contained in the dissemination.

## Acknowledgements

We are grateful to the Ministries of Health of Guinea, Liberia, Sierra Leone and Mali who permitted the conduct of the trial. We furthermore thank Alima, GOAL, and all site collaborators for their contribution in the implementation of original PREVAC trial. The authors and study team wish to thank the participants who consented to the trial.

