## Supplementary Appendix for "Cross-reactive Bundibugyo antibody responses after licensed Ebola vaccines"

**Cross-reactive antibody responses against Bundibugyo ebolavirus induced by licensed Ebola vaccines: analyses from the PREVAC randomized trial**

### **SECTION 1. PREVAC STUDY TEAM**

Jamila Aboulhab^2^ M.D., Michelle Aguirre-MacKenzie^12^ B.S., Pauline Akoo^3^ M.B., Ch.B., Esther Akpa^2^ M.S.N., M.P.H., R.N., Robert Akpata^1^ M.D., Sara Albert^17^ B.A., M.P.H, Boni Maxime Ale^4^ M.D., M.Sc., M.P.H., Serry Alimamy-Bangura^11^ M.B., Ch.B., Benetta C. Andrews^10^ M.D., Stephane Anoma^6^ M.D., Negin Atri^2^ M.P.H., C.P.H., Augustin Augier^6^ M.Com., Ken Awuondo^3^ M.Sc., Ahidjo Ayouba^30^ Ph.D., Moses Badio^10^ M.Sc., Aminata Bagayoko^1^ M.D., Abby Balde^17^ M.P.H., Joséphine Balssa^1,8^ Pharm.D., Lamin Molecule Bangura^11^ B.Sc., Kesha Barrington^17^ M.P.A., Eric Barte de Saint Fare^6^ B.A., Beth Baseler^17^ M.S., Ali Bauder^12^ B.A., P.M.P., Claire Bauduin^4^ M.Sc, Luke Bawo^10^ M.Sc., Abdoul Habib Beavogui^9^ M.D., Ph.D., Michael Belson^2^ B.S., Safaa Ben-Farhat^4^ M.Eng., Marion Bererd^6^ B.S., Nicolas Bernaud^1,18^ M.Sc., Teedoh Beyslow^10^ Pharm.D., Neirade Biai^30^ M.Sc., Jeanne Billioux^2^ M.D., Shere Billouin-Frazier^17^ M.Sc., Blandine Binachon^4^ M.D., M.P.H., Julie Blie^10^ M.Sc., Patricia Boison^17^ M.S., Fatorma Bolay^10^ Ph.D., Aliou Boly^6^ M.M., Rachael Elizabeth Bonawitz^12^ M.D., M.S., Anne-Gaëlle Borg^6^ M.C.M., Samuel Bosompem^2^ Pharm.D. M.Sc., Courtney Bozman^28^ M.Sc., Tyler Brady^2^ M.P.H., Sarah Browne^10^ R.N., B.S.N., Ryan Bullis^12^ Ph.D., Gabrielle Caberia^1^ Pharm.D. candidate, Barbara Cagniard^1^ Ph.D., Kelly Cahill^2^ R.N, M.Sc., C.C.R.C., R.A.C., Yingyun Cai^28^ Ph.D., Aissata Abdoulaye Camara^6^ M.Sc., Aboubacar Keira Camara^1^ M.D., M.Sc., Alseny Modet Camara^6^ M.D., Cécilia Campion^4^ M.Sc., Alexandre Cantan^4,29^ B.Sc., Jennifer Cash^17^ B.S., Michael Chea^10^ B.Sc., Geneviève Chêne^4^ M.D., Ph.D., Edward Choi^3^ Ph.D., Michelle Chouinard^6^ M.S.W., Florence Chung^1^ Ph.D., Lucy Chung^2^ Pharm.D., Papa Ndiaga Cisse^14^ Ph.D., Elfrida Cline-Cole^17^ M.A., Céline Colin^4^ M.Sc., Beth-Ann Coller^12^ Ph.D., Djélikan Siaka Conde^1^ M.D^.^, Katherine Cone^2^ M.S.W., LCSW-C, C-SWHC, Laurie Connor^12^ M.S., Nicholas Connor^3^ M.Sc., Joseph Boye Cooper^10^ M.Sc., Sandrine Couffin-Cadiergues^1^ Ph.D., , Mariam Coulibaly^20^ Pharm.D., M.Sc., Page Crew^2^ Pharm.D., M.P.H, B.C.P.S., Sandrine Dabakuyo-Yonli^4^ Pharm.D., Ph.D., Djeneba Dabitao^20^ Pharm.D., Ph.D., Bionca Davis^5^ M.P.H., Gibrilla Fadlu Deen^11^ M.D., M.Sc., Jean-François Delfraissy^1^ M.D., Ph.D., Christelle Delmas^1^ M.Sc., Mahamadou Diakite^20^ Pharm.D., D.Phil, Alpha Diallo^1,8^ M.D., M.P.H., Fatoumata Abdoulaye Diallo^6^, M.D., Mamadou Saliou Diallo^6^ M.D., M.P.H., Ayouba Diarra^20^ M.Sc., Samba Diarra^20^ M.Sc., Ph.D., Oualy Diawara^19^ B.S., Ilo Dicko ^20^ M.D, M.P.H., Bonnie Dighero-Kemp^28^ B.Sc., Samba Diop^21^ M.Sc., Ph.D., Waly Diouf^14^ Ph.D., Saurabh Dixit^2^ Ph.D., Barry Djenabou^6^ M.Sc., Laurie Doepel^2^ B.A., Eric D'Ortenzio^1,7,8^ M.D., M.P.H., Seydou Doumbia^20^ M.D., Ph.D., Moussa Moise Doumbia^19^ M.D., Nelson Dozier^28^ M.Sc., Natasha Dubois Cauwelaert^1,8^ Ph.D., Alain DuChêne^5^ B.S., Michael Duvenhage^17^ N.DIP.IT., Risa Eckes^2^ R.N., Elizabeth Elliott^2^ M.Sc., Luisa Enria^3^ Ph.D., Hélène Espérou^1^ M.D., Cécile Etienne^1^ M.Sc., Allison Eyler^17^ H.S.D., Lawrence Fakoli^10^ M.Sc., Mosoka Fallah^10^ Ph.D., Marie-Alix Fauvel^1^ M.Sc., Sylvain Faye^14^ Ph.D., John Fayiah^10^ M.Sc., Suzanne Fleck^3^ Ph.D., Vemy Fofana^6^ B.Comp, Karine Fouth Tchos^2^ M.D., M.P.H., Kokulo Franklin^10^ M.Phil., M.Sc., Daniela Fusco^1^ Ph.D., Auguste Gaddah^13^ Ph.D., Marylène Gaignet^1^ M.Sc., Katherine Gallagher^3^ Ph.D., Julie Gardner^12^, B.S., Harrison Gichini^28^ M.Sc., Julia Garcia Gozalbes^1^ M.D., Greg Grandits^5^ M.S, Maima Gray^10^ B.Pharm., Brian Greenwood^3^ M.D., Robin Gross^28^ M.Sc., Louis Grue^17^ R.N., B.S., B.S.N., Birgit Grund^27^ Ph.D., Oumar Guindo^20^ M.Sc., Pharm.D., Swati Gupta^12^ Dr.P.H., M.P.H., Fadima Haidara^19^ M.D., Benjamin Hamzé^1^ Pharm.D., Emma Hancox^3^ M.Sc., Patricia Hensley^3^ M.Ph., Lisa Hensley^28^ Ph.D., M.S.P.H., Betsey Herpin^2^, M.S.N, Elisabeth Higgs^2^ M.D., D.T.M.H., M.I.A., Trudi Hilton^3^ B.Pharm., M.Sc., Mickael Hneino^1^, Ph.D., Tracey-Ann Hoeltermann^28^, B.Sc., M.P.H., , Horace Preston Holley^17^ M.D., Marie Hoover16 Ph.D., Natasha Howard3 Ph.D., Melissa Hughes12 B.A., M.B.A., C.P.M., P.M.P., Sesay Idrissa^32^ B.Sc, Skip Irvine^12^ B.S., David Ishola^3^ M.D., Ph.D., Yvonne Jato^2^ M.P.H., Madison Joe^10^ M.Sc., Melvin Johnson^10^ M.Sc., Aboubacar Sidiki Kaba^6^ M.D., Jonathan Kagan^2^ Ph.D., Kade Kallon^17^, M.Sc., Michael Kamara^3^ MB.ChB, M.Sc., Myriam Kante^4^ B.S., Judith Katoudi6 M.D., M.P.H., Cheick Mohamed Keita^6^ M.D., Sakoba Keita^15^ M.D., Seykou Keita^22^ M.D., Stephen B. Kennedy^10^M.D., M.Sc., Babajide Keshinro^13^ M.B.B.S., F.W.A.C.P., Hassan Kiawu^10^ M.Sc., Mark Kieh^10^ M.D., M.S.M.H.C., Brent Killinger^12^ B.A., Moumouni Kinda^6^ M.D., M.B.A., Matthew Kirchoff^2^ Pharm.D., M.Sc., M.B.A., Gregory Kocher^28^ M.Sc., Mamoudou Kodio^19^ Pharm.D., Brian Kohn^3^ B.Sc., Lamine Koivogui^23^ Pharm.D., Ph.D., Richard Kojan^6^ M.D., Cece Francis Kolié^6^ Pharm.D., Jacques Seraphin Kolié^6^ M.D., David Kollie^10^ B.Sc., Stacy Kopka^17^ M.S., Bockarie Koroma^11^ B.Pharm., Dickens Kowuor^3^ B.Sc., M.Sc., Ph.D., Catherine Kpayieli-Freeman^10^ M.Sc., Ange-Marie D. Kpetigo^31^ M.Sc., Christine Lacabaratz^1,18^ Ph.D., Boris Lacarra^1^ M.D., Laurie Lambert^17^ B.S., Courtney Lambeth^12^ B.S., Solange Lancrey-javal^1,8^ Pharm.D., H. Clifford Lane^2^ M.D., Shadrach Langba^10^ B.Sc., Bolarinde Lawal^3^ M.Sc., Andrew Wen-Tseng Lee^12^ M.D., Shona Lee^3^ Ph.D., Shelley Lees^3^ Ph.D., Annabelle Lefevre^1^ M.D., Bailah Leigh^11^ M.D., M.Sc., Frederic Lemarcis^1^ Ph.D., Yves Lévy^1,18^ M.D., Ph.D., Edouard Lhomme^26^ M.D., Ph.D., Janie Liang^28^ M.Sc., Mameni Linga^10^ M.Sc., Ken Liu^12^ Ph.D., Brett Lowe^3^ M.Phil., Julia Lysander^10^ M.Sc., Ibrah Mahamadou^6^ Pharm.D., Irina Maljkovic-Berry^28^ Ph.D.*,* Marvington Mambiah^10^ A.Sc., Daniela Manno^3^ M.D., Ph.D., Jonathan Marchand^2,17^ M.S., Lindsay Marron^28^ M.Sc., Moses B.F. Massaquoi^10^ M.D., M.Sc., , Charly Matard^4^ B.S., Steven Mazur^28^ B.S., John McCullough^16^ B.S., Chelsea McLean^13^ Ph.D., Noémie Mercier^1^ Pharm.D., Pauline Michavila^6^ B.Bus., Tracey Miller^17^ R.N., B.S.N., Niouma Pascal Millimouno ^6^ M.D., Alejandra Miranda^17^ M.S., Soumaya Mohamed^6^ B.J., Tom Mooney^3^ B.A., Dally Muamba^6^ M.D., James Mulbah^11^ B.Pharm., Rita Lukoo Ndamenyaa^6^ M.D., M.Sc., James Neaton^5^ Ph.D., Désiré Neboua^1^ M.D., Micki Nelson^12^ B.S.N., M.S., Kevin Newell^17^ M.P.H., M.Ed., Vinh-kim Nguyen^24^ M.D., Yusupha Njie^3^ B.Sc., Wissedi Njoh^17^ M.S.N., Matthew Onorato^12^ B.S., Uma Onwuchekwa^22^ B.Sc., Susan Orsega^2^ M.S.N., FNP-BC, Inmaculada Ortega-Perez^1,8^ Ph.D., M.P.H., Cynthia Osborne^17^ B.S., Tuda Otieno^3^ M.SC., Davy Oulaï^4^ M.D., Sushma Patel^12^ M.S., P.M.P., Danielle Peart^2^ B.S., Martine Peeters^30^ Ph.D., James Pettitt^28^ M.Sc., Nathan Peiffer-Smadja^1^ M.D., Ph.D., Robert Phillips^3^ M.Sc., Jerome Pierson^2^ Ph.D., Peter Piot3 M.D., Ph.D., Micheal Piziali2 J.D., M.Sc., Stéphany Pong^1,8^ Pharm.D., Elena Postnikova^2^ Ph.D., Dudley Pratt^3^ M.D., Calvin Proffitt^17^ M.A., Alexandre Quach^1^ M.D., Sinead Quigley^1^ M.S.Sc., Nadeeka Randunu^2,17^ B.Sc., M.B.A., Laura Richert^26^ M.D., Ph.D., Priscille Rivière^1^ M.Sc., Céline Roy^4,29^, Ph.D., Amy Falk Russell^12^ M.S., Philip Sahr^10^ M.D., Katy Saliba^2^, M.Sc., Ph.D., Mohamed Samai^11^ M.B.B.S., Ph.D., Sibiry Samake^20^ Pharm.D., M.Sc., Jen Sandrus^17^ A.A., Ibrahim Sanogo^20^ Ms.P., M.D., Yeya Sadio Sarro^20^ Pharm.D., Ph.D., Serge Sawadogo^6^ M.D., M.Sc., Sani Sayadi^6^ M.D., M.P.H., Maxime Schvartz^1^ M.D., Christine Schwimmer^4^ Ph.D., Fatou Secka^3^ B.Sc., M.Sc., MB.ChB, Heema Sharma^28^M.Sc., Denise Shelley^17^ M.S., Bode Shobayo^10^ M.Sc., Sophia Siddiqui^2^ M.D., M.P.H., Jakub Simon^12^ M.D., Shelly Simpson^17^ M.S., Billy Muyisa Sivahera^6^ M.D., , Mary Smolskis^2^ B.S.N., M.A., Elizabeth Smout^3^ M.D., M.Sc., Emily Snowden^3^ M.A., Anne-Aygline Soutthiphong^4,29^ M.Sc., Amadou Sow^6^ M.Sc., Samba O. Sow^22^ M.D., M.Sc., Ydrissa Sow^2^ M.D., M.P.H., Michael Stirratt^25^ Ph.D., Léa Surugue^1^ M.J., Sienneh Tamba^10^ R.N., B.S.N., Cheick Tangara^20^ M.Sc., Milagritos D. Tapia^22^ M.D., Julius Teahton^10^ M.Sc., Jemee Tegli^10^ M.Sc., Monique Termote^4^ M.Sc., Guillaume Thaurignac^30^ M.Sc., Rodolphe Thiebaut^4^ M.D., Ph.D., Greg Thompson^5^ B.S., John Tierney^2^ B.S.N., M.P.M., Daniel Tindanbil^3^ M.Sc., Abdoulaye Touré^23^ Pharm.D., M.P.H., Ph.D., Elvis Towalid^10^ B.Pharm, Stacey Traina^12^ B.S., Awa Traore^19^ Pharm.D., Tijili Tyee^10^ Pharm.D., David Vallée^1^ Pharm.D., Renaud Vatrinet^1^ Ph.D., Corine Vincent^4^ M.Sc., Susan Vogel^2^ R.N, B.S.N., Cedrick Wallet^4^ M.Sc., Travis Warren^2^ Ph.D., Deborah Watson-Jones^3^ M.D., Ph.D., Wade Weaver^28^ M.Sc., Deborah Wentworth^5^ M.P.H., Cecelia Wesseh^10^ B.Sc., Hilary Whitworth^3^ Ph.D., Jimmy Whitworth^3^ Ph.D., Aurelie Wiedemann^1,18^ Ph.D., Wouter Willems^13^ Ph.D, Barthalomew Wilson^10^ M.Sc., Jayanthi Wolf^12^ Ph.D., Alie Wurie^11^ M.D., M.Sc., Delphine Yamadjako^17^ M.S., Marcel Yaradouno^6^ M.Sc., Quiawiah Yarmie^10^ M.Sc., Yazdan Yazdanpanah^1,7,8^ M.D., Ph.D., Shuiqing Yu^28^ B.S., Zara Zeggani^6^ M.Sc., Huanying Zhou^28^ B.S.

**Affiliations**

^1^ French Institute for Health and Medical Research (Inserm), 75013 Paris, France

^2^ National Institute of Allergy and Infectious Diseases, Bethesda, MD, USA or under contract/subcontract to NIAID

^3^ London School of Hygiene & Tropical Medicine, London, UK

^4^ Univ. Bordeaux, INSERM, Institut Bergonié, CHU de Bordeaux, CIC-EC 1401, Euclid/F-CRIN Clinical trials platform, F-33000 Bordeaux, France

^5^ School of Public Health, University of Minnesota, Minneapolis, MN, USA

^6^ The Alliance for International Medical Action, Alima, B.P.15530 Dakar, Sénégal

^7^ AP-HP, Hôpital Bichat-Claude Bernard, Service de Maladies Infectieuses et Tropicales, Paris F-75018, France

^8^ ANRS Emerging Infectious Diseases, Paris, France

^9^ Centre National de Formation et de Recherche en Santé Rurale de Maferinyah, Maferinyah, Guinea

^10^ Partnership for Research on Ebola Virus in Liberia (PREVAIL), Monrovia, Liberia

^11^ College of Medicine and Allied Health Sciences (COMAHS), University of Sierra Leone, Freetown, Sierra Leone

^12^ Merck Sharp & Dohme Corp, Inc., Kenilworth, NJ, USA

^13^ Janssen Vaccines and Prevention BV Leiden, The Netherlands

^14^ Département de Sociologie, FLSH, Université Cheikh Anta DIOP, Dakar, Sénégal

^15^ Agence Nationale de Sécurité Sanitaire, Conakry, Guinea

^16^ Advanced BioMedical Laboratories, L.L.C., 1605 Industrial Hwy, Cinnaminson, NJ, USA

^17^ Clinical Monitoring Research Program Directorate, Frederick National Laboratory for Cancer Research.

^18^ Vaccine Research Institute, Univ. Paris Est Créteil, Henri Mondor Hospital, Créteil, France

^19^ Centre pour le Développement des Vaccins, Ministère de la Santé, Bamako, Mali

^20^ University Clinical Research Center (UCRC), University of Sciences, Techniques and Technologies of Bamako (USTTB), Bamako, Mali

^21^ Liberia Institute for Biomedical Research Ethics Committee/National, Monrovia, Liberia

^22^ Center for Vaccine Development and Global Health, University of Maryland School of Medicine, 685 West Baltimore Street Baltimore, MD 21201-1509, USA

^23^ INSP (Institut Nationale de Santé Publique), Conakry, Guinea

^24^ École de santé publique de l’Université de Montréal, Montréal, Canada

^25^ National Institute of Mental Health, Bethesda, MD, USA

^26^ Univ. Bordeaux, INSERM, Institut Bergonié, CHU de Bordeaux, CIC-EC 1401, Euclid/F-CRIN clinical trials platform and U1219 BPH Inria Sistm, F-33000 Bordeaux, France

^27^ School of Statistics, University of Minnesota, Minneapolis, MN, USA

^28^ Integrated Research Facility at Fort Detrick (IRF-Frederick), National Institute of Allergy and Infectious Diseases, National Institutes of Health (NIH), Fort Detrick, Frederick, MD, USA

^29^ Univ. Bordeaux, INSERM, MART, UMS 54, F-33000 Bordeaux, France

^30^ Recherche Translationnelle Appliquée au VIH et aux Maladies Infectieuses, University of Montpellier, Institut de Recherche pour le Développement, INSERM, 34090, Montpellier, France

31 Bordeaux Population Health Research Centre, Université de Bordeaux, Inserm, and Inria, Bordeaux, France
32 University of Management and Technology UNIMTECH, FreeTown, Sierra Leone.

### **SECTION 2. RESULTS**

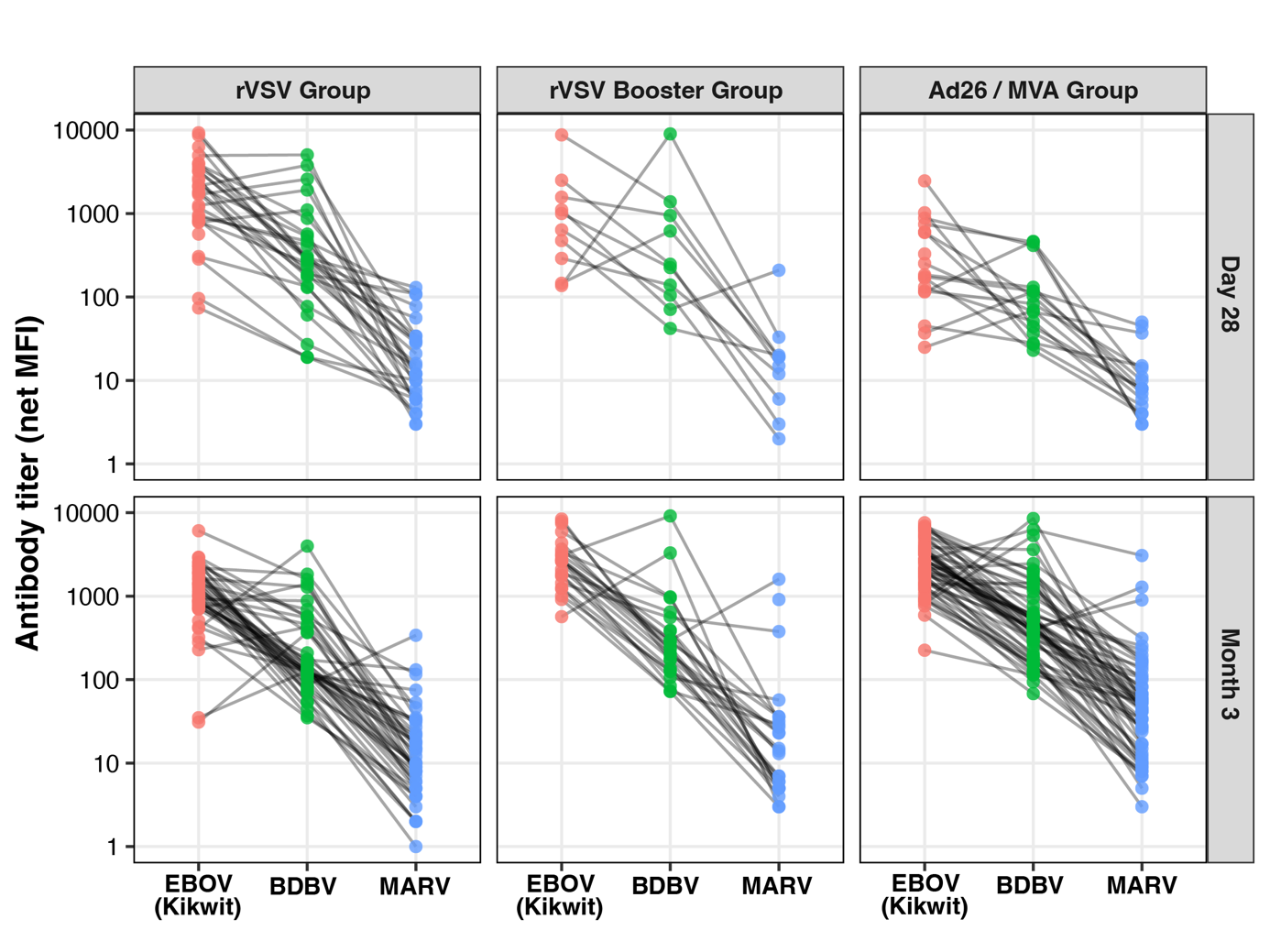

#### **Figure S1. Description of the binding antibody response measured by multiplex Luminex (net MFI) at Day 28 and Month 3 after rVSV, rVSV booster and Ad26 / MVA vaccination, respectively.**

Paired antibody responses against Kikwit Ebolavirus (EBOV), Bundibugyo virus (BDBV) and Marburg virus (MARV) glycoproteins for individual participants. Each line represents paired measurements from the same participant.
